# Large-scale Genomic Survey of Non-typhoidal *Salmonella enterica* serovar Minnesota Strains in Chicken Products Reveals the Emergence of Multidrug Resistant Clones

**DOI:** 10.1101/2024.07.28.24311129

**Authors:** Jiayi Huang, Khaloud O. Alzahrani, Ge Zhou, Shahad A. Alsalman, Ayidh M. Almansour, Meshari A. Alhadlaq, Shaykhah Alhussain, Abdullah A. Alajlan, Saleh I. Al-Akeel, Malfi S. Al Rashidy, Abdulrahman Alzauhair, Fahad M. AlReshoodi, Amani T. Alsufyani, Nourah M. Alotaibi, Afnan Althubaiti, Elaf Alshdokhi, Ashwaq Alhamed, Manal Almusa, Talah Almadi, Nouf Almutairi, Lenah E. Mukhtar, Abdulmohsen L. AlHarbi, Manuel Banzhaf, Mathew Milner, Mohammad AlArawi, Sulaiman Alajel, Danesh Moradigaravand

## Abstract

**Background:** *Salmonella enterica* serovar Minnesota (*S*. Minnesota) is an emerging serovar of non-typhoidal *Salmonella*, known to persist in the food chain and distribution systems, potentially leading to outbreaks of *Salmonella* infections in human settings. Understanding the population genomics and dynamics of this pathogen is key to designing preventative measures and containing its spread within the poultry production chain.

**Methods:** In this study, we conducted a large-scale study on *S*. Minnesota outbreak by fully characterizing population diversity and dynamics of a systematic collection from the poultry production chain in the Kingdom of Saudi Arabia (KSA). We sequenced 260 *S*. Minnesota strains from the western, eastern, and central regions of the country. We analyzed sequencing data to decipher the population diversity and dynamics of multidrug resistant strains and characterize the genetic basis of resistance and virulence. A hybrid long- and short-read sequencing approach was employed to analyze the population diversity of plasmids carrying antimicrobial resistance and virulence factors.

**Results:** Our results indicate the rise of four clones (Bayesian Analysis of Population Structure; BAPS groups) in Saudi Arabia, three of which were mixed with global strains. The clones emerged over the past five to ten years and exhibited circulation between countries. The transmission analysis shows evidence of the spread of strains across cities, between countries, and mixing of strains from different suppliers, on epidemiological time scales. The emerging clones also harbored a higher resistance and virulence level than ancestral clones, owing to the acquisition of multiple plasmids, most importantly the IncC plasmid. The IncC plasmid was a mosaic plasmid, which carried antimicrobial resistance islands with *bla*_CMY-2_, ESBL *bla*_CTX-M_, aminoglycoside, and tetracycline resistance genes, as well as a hyperpathogenicity island with yersiniabactin genes. The plasmidome analysis revealed a high level of dynamics in the IncC plasmid structures with various configurations of resistance genes.

**Conclusion:** Taken together, our results demonstrate a dynamic population and the emergence of multidrug-resistant clones in *S*. Minnesota. The results also highlight the role of plasmid acquisition and genomic variations in driving the concurrent evolution of pathogenicity and resistance in *S*. Minnesota.

## Introduction

Non-typhoidal *Salmonella* (NTS) is recognized as the predominant foodborne bacterial pathogen. Each year, foodborne NTS is responsible for an estimated 93 million cases of gastroenteritis and 155,000 deaths worldwide ^1^, contributing to approximately 4 million disability-adjusted life years ^2^. In resource-limited regions and among immunocompromised individuals, extraintestinal or invasive NTS infections, often linked to specific serovars or phylogenetic clades, significantly contribute to the burden of the disease. Invasive cases can progress to life-threatening bloodstream infections, particularly problematic due to the increasing prevalence of antimicrobial resistance (AMR) in NTS ^3^. This resistance can spread directly through human contact with AMR bacteria from food animals or indirectly via resistant bacteria contaminating environmental reservoirs such as water and soil, driven by the use of antibiotics in agriculture ^4^. Assessments of NTS antimicrobial susceptibility patterns over time indicate a concerning trend of moderate to high levels of resistance, including multidrug resistance ^5^.

*Salmonella* serotype Minnesota (*S.* Minnesota) has recently emerged as a notable NTS serovar, exhibiting increased prevalence within poultry farming and related food products, such as chicken meat and eggs, in recent years. The prevalence of *S.* Minnesota rose following the introduction of a vaccine targeting *Salmonella enterica* serovar Enteritidis in 2003, which was the predominant Salmonella serovar in Brazilian poultry and exported chicken meat at that time ^6,7^. Alongside vaccination, widespread antimicrobial use in animals is believed to have influenced these shifts ^8^. The increased worldwide demand for animal protein is expected to double the antimicrobial usage in livestock in Brazil by 2030 ^9,10^. Since Brazil serves as a major exporter of poultry to countries worldwide, numerous reports highlight the global dissemination and spread of *S.* Minnesota throughout food export chains, a process occurring over extended epidemiological timescales.

The pathogenic mechanisms of *S.* Minnesota remain largely unknown, primarily due to its rarity and the limited number of studies focused on this serotype. Despite its infrequent isolation, several significant cases have been documented. First reported in the 1950s, a major outbreak of food poisoning resulted in over 500 clinical cases linked to the consumption of meat pies produced in a factory in South-West Northamptonshire. The investigation revealed that the meat pies were the vehicle of infection ^11^. Since this initial report, isolations of *S.* Minnesota from clinical specimens have been infrequent. In 1985, an acute gastroenteritis outbreak occurred in a camp in Saudi Arabia ^12^. Another notable case involved a patient with Crohn’s disease who developed urosepsis due to *S.* Minnesota without typical gastrointestinal symptoms. This case highlighted the necessity of considering NTS in immunocompromised patients presenting with severe infections ^13^.

Whole genome sequencing, which provides a comprehensive view of genetic biomarkers, has been increasingly used in food surveillance and monitoring of bacterial contaminations ^14^. Few studies have employed the fine resolution of whole genome sequencing to elucidate the diversity and genetic profiles of antimicrobial resistance in *S.* Minnesota. These studies have shown the persistence of *S.* Minnesota across food chains in various countries and the emergence of clones over the past few decades ^10^. Furthermore, these studies have highlighted the role of IncI1 or IncC (formerly known as IncA/C ^15^) plasmids as the main drivers of resistance genes against third-generation cephalosporins, tetracyclines, and/or sulphonamides by carrying respective genes such as *bla*_CMY-2_, *tet*(A) and *sul2* ^16–18^ ^19–21^. Some studies have also reported the occurrence of *mcr* genes, which confer resistance against the last-resort antimicrobial colistin, suggesting an ongoing rise in resistance levels for clinically important antimicrobials ^22,23^. Despite these significant insights, studies conducted at the country level often had limited sample sizes, making it difficult to capture the population dynamics of outbreaks and the regional epidemiology of multidrug resistant clones in *S.* Minnesota.

In this study, we present large-scale population genomics insights into the diversity and dynamics of *S.* Minnesota within the chicken production chain in a nationwide study across Saudi Arabia, a country known for its high poultry production and consumption rates. The poultry industry in Saudi Arabia has undergone significant growth in recent years. Poultry is a staple protein source in Saudi Arabia, with individuals consuming an average of 47 kg of chicken meat and 120 eggs per year ^24^. In 2020 alone, the country produced 900,000 metric tons of chicken meat, satisfying 60% of domestic demand, and is projected to grow at a compound annual growth rate of 3.47% from 2020 to 2025 ^25^. During the same year, Saudi Arabia imported a total of 652,283 metric tons of chicken meat and related products, with Brazil being the primary supplier, contributing 72% (467,522 mt) of these imports ^26^ ^27^. Clinically, Salmonella infection (salmonellosis) is one of the most frequently reported bacterial infectious diseases, with around 2,000 cases annually ^28,29^. The depth and breadth of the collection allowed us to fully elucidate the population dynamics of *S.* Minnesota clones within the country and identify key factors driving the emergence and spread of antimicrobial-resistant clones.

## Materials and methods

### Ethical Approval

The study was approved by the Institutional Review Board (IRB) of the General Administration of Research and Studies, Ministry of Health, Saudi Arabia (No. 23-23 M), and the Institutional Biosafety and Bioethics Committee (IBEC) of King Abdullah University of Science and Technology (No. 22IBEC046).

### Sample collection and isolation

Between 2020 and 2022, a total of 260 Salmonella Minnesota isolates were collected with the facilitation of the Saudi Food and Drug Authority (SFDA). This collection was part of multiple monitoring and surveillance programs adhering to the sampling guidelines outlined in the document “Integrated Surveillance of Antimicrobial Resistance in Foodborne Bacteria” by the World Health Organization. ^30^. Among these, 173 isolates originated from domestically produced poultry samples obtained from various supermarkets across Saudi Arabia, while the remaining 87 isolates were sourced from imported poultry products from Brazil. Isolates were retrieved from 17 suppliers (brand code). This study also included a single *S.* Minnesota isolate recovered from a patient admitted with clinical Salmonellosis to a hospital in Jeddah, Saudi Arabia. Informed consent was obtained from the participant prior to sample collection.

Isolation of *Salmonella* from chicken meat samples was carried out according to ISO 6579-1:2017/Amd 1:2020 (International Organization for Standardization, Geneva, Switzerland) ^31^. Upon arrival at RLM, all isolates were further confirmed at the species level using Matrix-assisted laser desorption/ionization-time of flight (MALDI-TOF). Further serological testing to determine their serotypes was conducted according to the Kauffman-White scheme using slide agglutination tests with commercially available mono- and poly-O group Salmonella A, B, C, D, E antisera (Remel, Europe Ltd., United Kingdom). Isolates were also subjected to *Salmonella* real-time PCR 7500 using the MicroSEQ™ Salmonella spp. detection kit (Thermo Fisher, United States) in food. The information on the isolates source, type and date of collection is provided in Supplemental Table S1.

### Antimicrobial susceptibility testing

The antimicrobial susceptibility testing (AST) for all *S.* Minnesota isolates was conducted using the microbroth dilution (MBD) method with the Sensititre EUVSEC3 panel manufactured by TREK Diagnostic Systems (Thermo Fisher Scientific). Inoculation and incubation procedures followed the manufacturer’s guidelines. The panel included 15 antimicrobials from various classes: aminoglycosides (amikacin, gentamicin), penicillins (ampicillin), macrolides (azithromycin), cephalosporins (cefotaxime, ceftazidime), carbapenems (meropenem), quinolones (ciprofloxacin, nalidixic acid), polymyxins (colistin), phenicols (chloramphenicol), tetracyclines (tetracycline), glycylcyclines (tigecycline), and folate pathway inhibitors (trimethoprim, sulfamethoxazole). These antibiotics are included in European antimicrobial surveillance programs, particularly the European Committee on Antimicrobial Susceptibility Testing (EUCAST), which monitors resistance patterns to inform treatment guidelines and public health policies. *Escherichia coli* ATCC 25922 served as the quality control reference strain. Minimum inhibitory concentrations (MICs) for each antimicrobial were interpreted according to Clinical and Laboratory Standards Institute (CLSI) guidelines ^32^. For streptomycin, azithromycin, and cefoxitin, National Antimicrobial Resistance Monitoring System (NARMS) guidelines were applied due to the absence of CLSI interpretative breakpoints. Tigecycline interpretation followed FDA criteria, as no CLSI or NARMS breakpoints were available ^33^.

### DNA extraction and Short-read sequencing

Genomic DNA was extracted using the DNeasy Power Soil kit following the manufacturer’s instructions (Qiagen, CA, United States). The purity of genomic DNA was assessed by measuring the A260/A280 ratio (target ≥1.8) using the QIAxpert system (Qiagen, CA, United States), and DNA concentration was quantified with the Qubit system (Qubit™ fluorometer, Invitrogen, CA, USA) according to the manufacturer’s protocol. Libraries were prepared using the Nextera XT DNA sample preparation kit and the Nextera XT Index Kits (Illumina, Inc., San Diego, CA, United States). Following index PCR, samples were purified with 45 μl of Agencourt AMPure XP magnetic beads at a sample to beads ratio of 3:2 (Beckman Coulter, Brea, CA, United States), normalized based on Qubit quantification, and pooled. Subsequently, libraries were sequenced in a 250 bp paired-end-read format using the Illumina NovaSeq 6000 kit with 5% PhiX control (Illumina Inc., San Diego, CA, United States), following the manufacturer’s instructions.

### Short-read sequencing data analysis

Sequence read quality control and assembly quality assessment were conducted using fastQC v.0.11.3 (http://www.bioinformatics.babraham.ac.uk/projects/fastqc/) and Trimmomatic v.0.36 software. Short reads underwent *de novo* assembly using the Unicycler v0.5.0 pipeline (https://github.com/rrwick/Unicycler) with default parameters. Contigs shorter than 200 bp were excluded from the assemblies. We employed srst2 v0.2.0 (www.github.com/katholt/srst2) ^34^ with a sequence similarity and coverage threshold of 90% to analyze assemblies against the PlasmidFinder (v2.1.1) ^35^, VFDB (v6.0) ^36^, and CARD (v3.2.8) ^37^ databases for detecting plasmid replicons, resistance and virulence genes, respectively. Resistance genes were confirmed with AMRFinderPlus (V.3.11.11)^38^. We evaluated the odds ratio of resistance phenotype associated with the presence of each resistance gene using the odds-ratio function from the Epitools package (v0.5.10.1) in R. Resistance mutations were identified using staramr v0.7.1 (https://github.com/phac-nml/staramr). To reconstruct the pangenome, assemblies were annotated using Prokka v1.14.5 ^39^. The annotated assemblies were then analyzed using Panaroo with default parameters ^40^. For phylogenetic analysis, reads were aligned to the reference genome of *S.* Minnesota (accession number: CP017720.1) using the Snippy pipeline (https://github.com/tseemann/snippy) with default settings. The FastTree package (2.1) was used with default parameters to generate an approximately maximum likelihood phylogenetic tree ^41^.

### Contextualization of the samples

To contextualize our collection, we retrieved all available genomic data as of February 21, 2024, from EnteroBase (www.enterobase.warwick.ac.uk) ^42^ and Pathogen Detection Databases (www.ncbi.nlm.nih.gov/pathogens). After removing duplicates and excluding data with insufficient metadata regarding country and date of isolation, we curated a dataset comprising short-read sequencing data from 507 isolates. These short-read sequences were aligned to the reference genome as previously described. We calculated pairwise Single Nucleotide Polymorphism (SNP) distances between strains and performed hierarchical clustering using the BAPS method ^43^, as implemented in R ^44^, with one round of iteration to identify clusters within the dataset (Isolates with dates of isolation were selected). We reconstructed the minimum spanning tree for the BAPS groups by first computing the average pairwise SNP distance between genomes in different BAPS groups and then using the average distance matrix between BAPS groups as input for the neighbor-joining tree function in the ape package (V.5.0) ^45^. The information of the retrieved genomes is provided in Supplemental Table S1.

### Phylodynamics analysis

We conducted a phylodynamic analysis to estimate the epidemiological parameters for the BAPS groups containing Saudi isolates (BAPS groups 1-4). Initially, we extracted Saudi genomes and mapped the short reads against a local reference genome for each BAPS group. The local reference genome was constructed by concatenating the genomic contigs of the genome with the best assembly statistics (i.e., highest N50). After mapping the short reads, we extracted SNP sites from the multiple alignment of genomes and removed hypervariable regions using Gubbins ^46^ with five iterations.

The resulting alignment was used for phylogeographic diffusion analysis with BEAST (v2.0) [45]. We employed the built-in MASCOT (Marginal Approximation of the Structured Coalescent) approach, which is integrated into the BEAST package and uses a structured coalescent approximation ^47,48^ to infer ancestral migration histories. This method is commonly applied to quantify the geographic spread of pathogens between cities by leveraging both sample locations and sequence data. We used the city of isolation as a discrete state for each taxon. For the prior model, we used a constant population size model with a uniform distribution for the clock rate. We opted for a symmetric model with a uniform distribution for the discrete trait (i.e., city of isolation) ^49^.

We ran the Markov chain Monte Carlo (MCMC) chains for 10^8^ iterations and assessed the convergence of the chains by ensuring that the effective sample size (ESS) for key parameters exceeded 200. Convergence was achieved for the BAPS2 and BAPS4 clusters, but not for BAPS3. We removed the first 10% of trees as the burn-in. The remaining trees were merged using the TreeAnnotator tool included in the package, and the mean and 95% highest posterior density (HPD) intervals for key epidemiological parameters were extracted from the output file. To study the global spread of BAPS2 and BAPS4 clones, we repeated the above analysis using the country of isolation as the geographical state instead of cities after including genomes from other countries (external) in the same BAPS group to the internal genomes.

### Transmission analysis

We reconstructed networks of potential recent transmission events among strains within each BAPS group using the R package adegenet (V.1.3-1) ^50^. The tool inferred maximum likelihood networks based on pairwise SNP distances between samples, considering their isolation dates. To compute these distances, we took the SNP alignment for each BAPS groups after removing hypervariable sites, as detailed above. Subsequently, we utilized the distdna tool in the ape package in R with default parameters for each BAPS group. We then fed the distance matrix and dates of isolations into the seqTrack function in the adegenet package. For defining the networks, we applied a cutoff of 10 SNPs, which, based on the estimated recent substitution rate of the core genome of *S.* Minnesota (detailed in the Results), corresponds to SNPs accumulated over approximately 5 years. The resulting networks were visualized using the igraph library in R.

### Long-read sequencing and plasmidome analysis

Plasmid DNA was extracted from a 4 mL overnight LB culture using the QIAprep® Miniprep kit according to the manufacturer’s instructions (Qiagen, CA, United States). The concentration and purity of the plasmid were determined using Qubit HS dsDNA kits and spectrophotometry at 260/280 nm. Libraries for sequencing various plasmid isolates were prepared, multiplexed using 96-plex Rabid Barcoding Kits, and loaded onto MinION flow cells (Oxford Nanopore Technologies) for a 48-hour run following the manufacturer’s protocol. Subsequently, hybrid assembly was performed with Unicycler using the conservative mode to reconstruct the chromosome and plasmid genomic fragments, which were examined and verified using Bandage (v0.9.0) [48]. Annotation of the assemblies to identify potential plasmids was conducted using Prokka, and the plasmid pan-genome was reconstructed using Panaroo ^40^. To visualize the network, Cytoscape (v3.10.1) was employed (available at https://cytoscape.org). Plasmids were visualized and annotated using the Proksee portal and its built-in tools (www.proksee.ca) ^51^. Gene-level alignment and visualizations were performed using the pyGenomeViz (v1.0.0) package in Python (https://github.com/moshi4/pyGenomeViz). The assembled genomic fragments for the plasmids were then analyzed for the presence of complete origin of replication sequences, virulence factor genes, and antimicrobial resistance genes with blast searches against the aforementioned databases.

We extracted all plasmid genomic fragments containing a known plasmid replicon and then computed sequence similarity using an alignment-free approach. Initially, we generated k-mers of size 50 for each plasmid. Subsequently, we constructed a distance matrix and computed pairwise Jaccard distances, defined as 1 - |intersection of kmers|⁄|union of kmers|, between all plasmid fragments. Using fviz_nbclust in R, we employed k-means clustering and identified five clusters as the optimal number, i.e. the number that maximized the within-cluster sum of squares, via the elbow method. Upon further inspection, two highly similar plasmid sequence clusters were merged to yield final four clusters, which were characterized. To obtain the phylogenetic distribution of plasmid in the entire *S.* Minnesota population, we mapped the short reads for the strains against the representative plasmids from each cluster using Snippy, as detailed above, and used the cut-off of 90% for detection of the plasmid. For plasmid annotation, we searched the NCBI nucleotide BLAST database for similar sequences in the public repository that matched our plasmids.

### Availability of data

The short- and long-read sequencing data were submitted to the European Nucleotide Archive (ENA) under study number PRJEB67737. Metadata, profiles of antimicrobial resistance genes/mutations, virulence factor genes, and plasmid replicons are provided in Supplemental Table S1. All other intermediate files and relevant tables, e.g. clustering information of plasmids and plasmid sequences, are available on the GitHub directory for the project: https://github.com/hjy1805/Sminnesota_project.

## Results

### Global population structure indicates recent emergence and expansion of clones

Over the course of 18 months, we screened chicken products, including processed and unprocessed meat, and identified 260 samples for whole genome sequencing. Among these, 87 strains were of Brazilian origin, while the rest were isolated from local Saudi food products (Supplemental Table S1). To contextualize the genomes, we also retrieved genomes with available data from public repositories of global collections (Figure S1A). The population structure consisting of isolates sequenced in the current study (internal isolates) and external genomes revealed 14 clones, eleven of which consisted of more than ten isolates, ranging from 20 to 140 genomes. We designated these as BAPS groups BAPS1-11 (Figure S1A). Except for two BAPS groups (BAPS1 and BAPS4), the other groups included strains from human sources (Figure 1A, Figure 1). Isolates from different continents were unevenly distributed across the major clones. Four clusters, namely BAPS10, BAPS11 and BAPS9 consisted exclusively of strains from America and Europe (Figure S1A, Figure 1). Internal genomes fell into six BAPS groups, specifically BAPS groups 1 to 6 (Figure 1A, Figure 1). BAPS1 and BAPS6 clusters included Saudi and Brazilian isolates, respectively. BAPS1 to BAPS6 groups exhibited lower genetic diversity and included more recent isolates (p-value from Wilcoxon signed-rank test < 0.05), clustering together (Figure 1B, Figure 1). Brazilian samples co-occurred in BAPS groups 2 to 4 with Saudi samples. The phylogenetic tree, reconstructed from the average genetic distance of the genomes in different BAPS groups, revealed that clones with Saudi isolates formed recently emerging clones, directly descended from the older BAPS6 group, which included Brazilian and European samples and descended from older North American and European BAPS clones. This finding aligns with the Brazilian origin of imported meat to Saudi Arabia (Figure 1). The genome of the only human sample recovered from a salmonellosis infection in our study, K_287, showed genetic identity with the food samples and grouped with genomes in BAPS2. The most closely related genomes were K_099 and K_258, recovered from fresh chicken in Dammam (11 and 14 SNPs distant, respectively), and K_017 from fresh chicken in Riyadh (19 SNPs distant), all sharing the same brand code (supplier). This suggests potential routes of bacterial food contamination into human settings.

**Figure 1:**
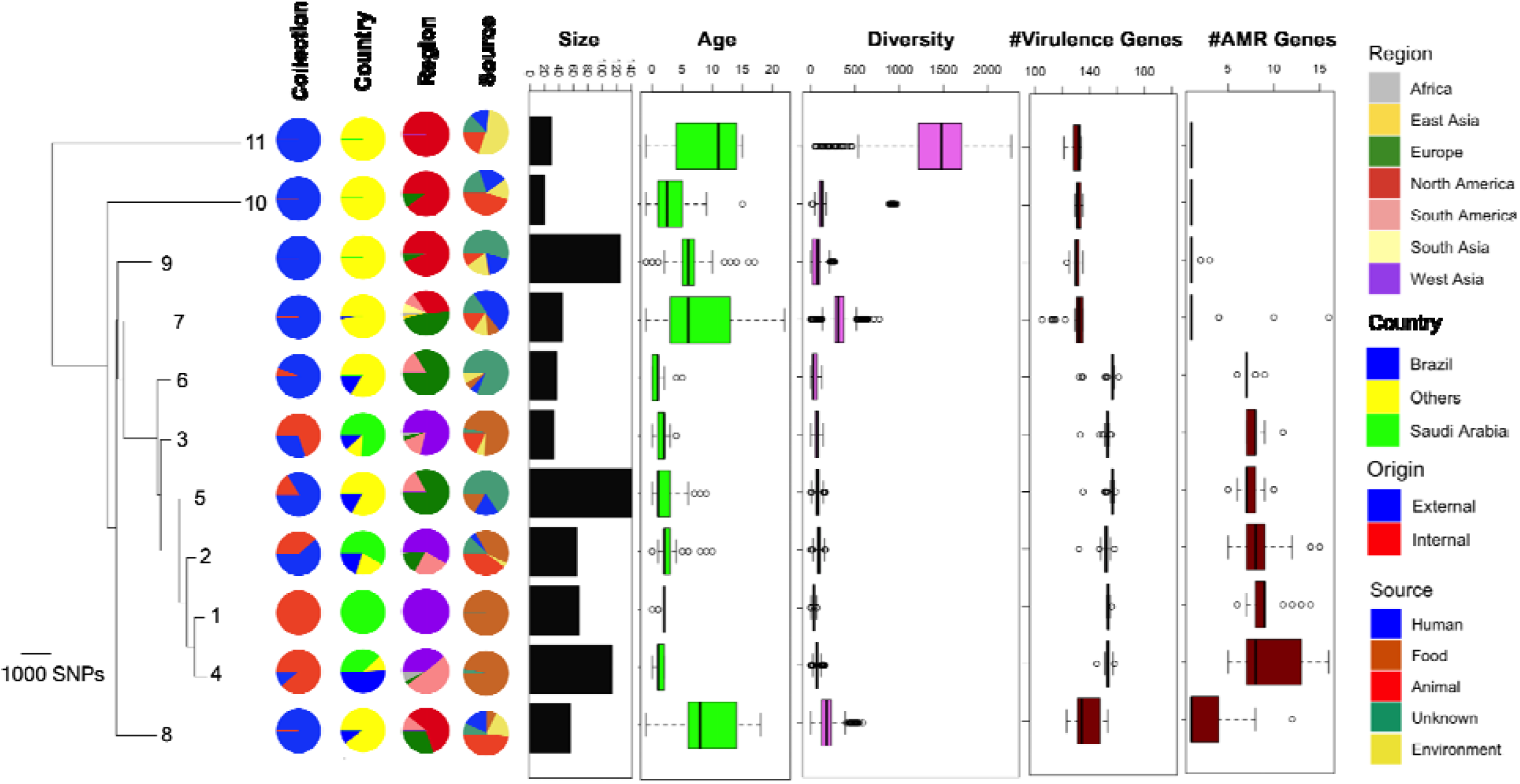
Population structure of major clones in the *S.* Minnesota population. The tree is a minimum spanning tree reconstructed from the mean SNP distances between pairs of isolates in different BAPS groups shown in Figure 1, and it is midpoint-rooted. The boxplot for diversity corresponds to pairwise SNP distances between genomes within each BAPS group. The boxplots for the count (#) of virulence and resistance genes show genes identified by the SRST2 pipeline. Internal genomes are those sequenced as part of this study, while external genomes are from public databases (EnteroBase/PathogenDetection). The boxplots for age show the distribution of the years of collection for the strains in each BAPS group.

### Recent clones concurrently emerged and expanded clones on country and global level

Next, we examined the dynamics of clonal expansion across the country (BAPS1 to 4) (Figure 2A). All clones formed concurrently over the course of 5 to 7.5 years (Figure 2A). The inferred substitution rates for the clones averaged between 7.5 × 10^−7^ and 2.5 × 10^−6^ substitutions per site per year, which overlaps with the mutation rates of *Salmonella enterica* serovar Typhimurium, ranging from 1.9 × 10^−7^ to 1.49 × 10^−6^ substitutions per site per year ^52–54^ (Figure 2A). All clones exhibited a growing population trend since their formation, with slight variations. For BAPS1, BAPS3, and BAPS4, the population growth either slowed down or reversed recently, while for the most recent BAPS2 clone, the population continued to expand (Figure 2B). The clone also exhibited other signatures of rapid expansion, characterized by the relatively highest effective population size and the highest substitution rate (Figure 2A). We extended the analysis for BAPS2 and BAPS4 clones to include genomes from other countries. Phylodynamic trees confirmed the formation of Saudi clones between 2010 and 2014 (Figure 2C). In the BAPS2 group, Brazilian samples served as ancestors to the Saudi clone and other clones in the UK and South Africa. The Saudi clade served as the ancestor for a lineage from the UAE ^27^, a country that imports poultry meat from Saudi Arabia, demonstrating the global and regional distribution of the clone (Figure 2C). The BAPS4 tree indicates the concurrent expansion of two Saudi clades: one originating from a Brazilian clade in 2011 (95% HPD: 2010-2014) and the other from a Portuguese lineage in 2008 (95% HPD: 2006-2011) (Figure 2C). The Brazilian clade gave rise to multiple lineages in the UK, South Africa, and Germany, illustrating the global circulation of the clones.

**Figure 2:**
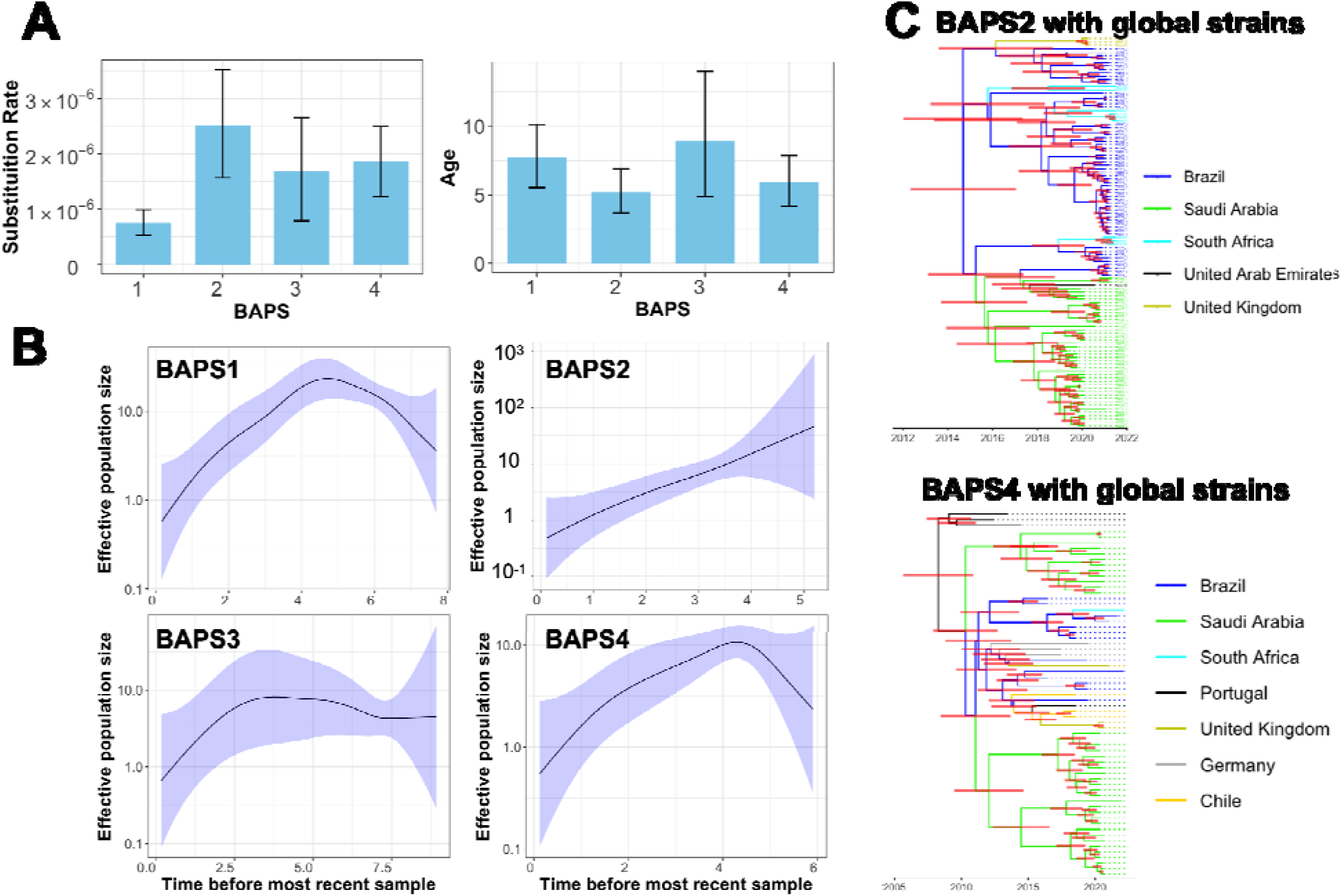
Phylodynamic analysis of the clones circulating in Saudi Arabia. **A)** Estimated clone age for four clones in the population. Error bars represent the 95% highest posterior density (HPD). **B)** Skyline growth plot depicting changes in population size over time. The shaded region indicates the 95% confidence interval. **C)** Phylodynamic trees for BAPS2 and BAPS4 groups, including genomes from the global collection. Internal branch colors denote the inferred status of the country of origin, with horizontal red bars indicating the 95% HPD.

### Global and local transmission patterns of *S.* Minnesota

We further examined the genetic relatedness between cities by inferring ancestry relationships among the sampled sequences. We therefore reconstructed genealogies of sampled sequences under the assumption that ancestors and descendants were sampled together. By using a sequence identity cutoff of 10 SNPs, which corresponds to transmission within approximately 2 years, we reconstructed the maximum parsimony genealogy for genomes within each BAPS clone (Figure 3). For BAPS groups 1 to 4, we identified 7, 4, 2, and 9 transmission sub-networks, respectively (Figure 3). Most sub-networks comprised samples from different cities or countries, illustrating the extent of global and local spread of the strains. Specifically, the mixing of samples within the same city from different suppliers was noted in the BAPS1 group (Figure 3). At the international level, BAPS3 and BAPS4 included networks with samples from South Africa/Portugal and South Africa/Brazil, respectively. In the BAPS4 group, Brazilian samples, retrieved as part of the current study, served as ancestors for the Saudi samples in 7 out of 9 sub-networks, likely through single or multiple import events (Figure 3). These findings confirm the results of the phylodynamic analysis, highlighting the simultaneous local and global circulation of the strains within the food chain across cities and countries. This pattern is also consistent with the dual role of poultry industry of Saudi Arabia as both an importer/exporter of poultry and a site of local production.

**Figure 3:**
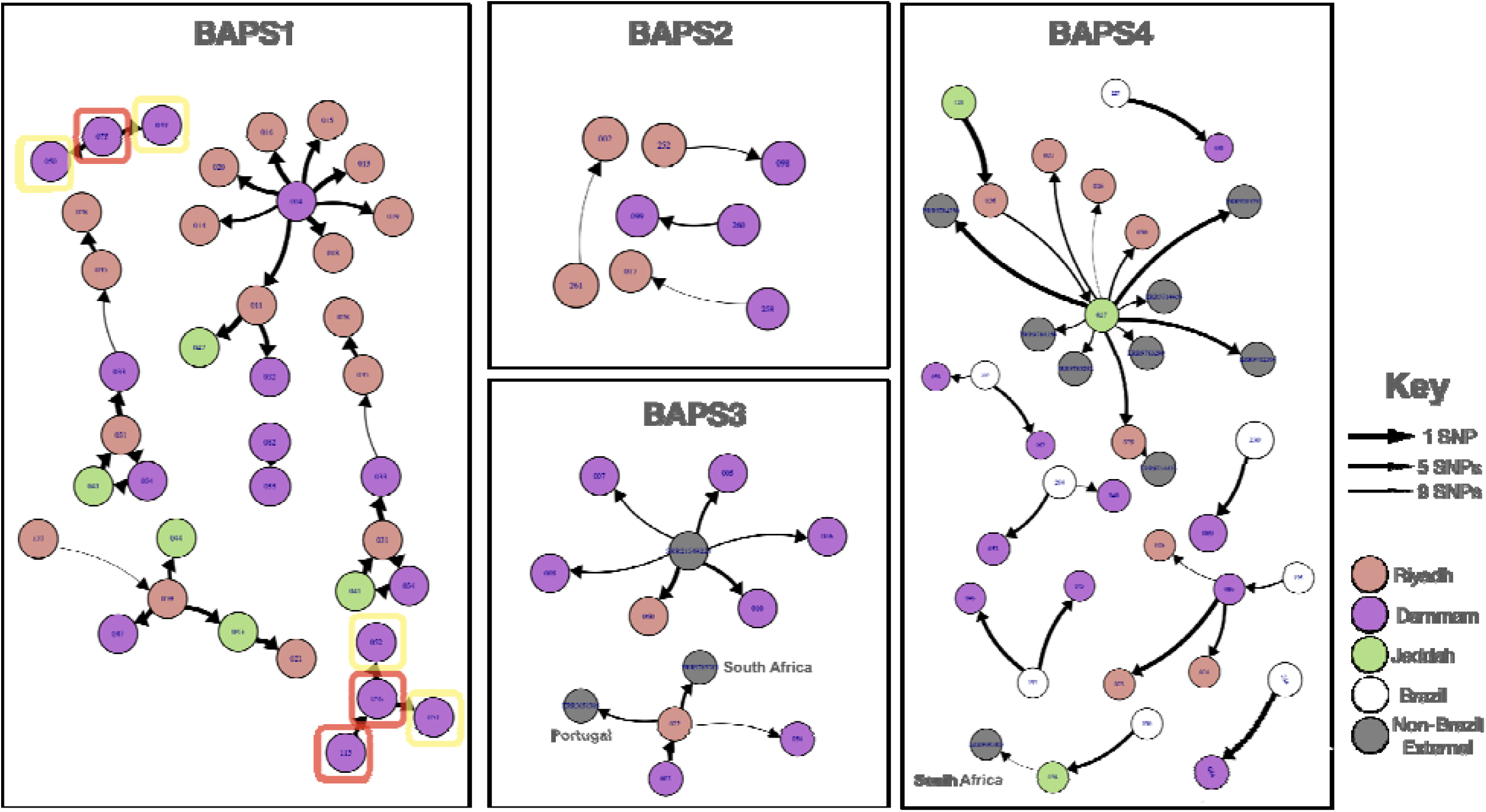
Transmission network for the four BAPS groups. Each node represents one genome, and edges indicate genetic relatedness, with a distance cutoff of 10 defining the networks. External (global) isolates are colored grey. All external isolates in the BAPS4 group originated from South Africa. Colored squares denote different brand codes/suppliers for isolates from the same city. Each color represents a distinct brand code/supplier. White nodes within BAPS4 represent Brazilian samples from the current study.

### Higher frequency antimicrobial resistance and virulence gene in recent *S.* Minnesota clones

Despite lower genetic diversity, clones with Saudi isolates (BAPS1 to 5) and the clone with internal Brazilian samples (BAPS6) exhibited a significantly higher number of antimicrobial resistance and virulence factor genes compared to older clones (p-value from Wilcoxon signed-rank test < 0.05) (Figure 1). For BAPS1 to 6, strains harbored an average of 7.5 (95% CI: 5.7-9.4) resistance genes, while for older BAPS clones, the count was 0.5 (95% CI: 0.1-2.4). Similarly, for BAPS1 to 6, strains harbored an average of 148 (95% CI: 142-155) unique virulence genes, whereas for older BAPS clones, the count was 132 (95% CI: 123-138). The estimated rates of acquisition of resistance and virulence genes over time were 0.22 (95% CI: 0.12-0.34) and 12.4 (95% CI: 11.3-13.8), respectively, indicating increasing virulence and resistance levels in recent *S.* Minnesota clones.

### Diversity of genes and genomic backgrounds for antimicrobial resistance genes

The collection from S. Minnesota exhibited a range of acquired genes conferring resistance to various antimicrobials. These included aminoglycosides (*aac6’, aac3,ant3’’, aph3’, aadA* and *aph4*), β-lactams (*bla*_TEM_ *bla*_CTX-M_ and *bla*_CMY-2_), tetracyclines (*tetA*, *tetB* and *tetD*), polymyxins (*mcr1* and *mcr9*), florfenicol (*floR* and *cmlA*), macrolides (*cat, lnuF* and *mphA*), fluoroquinolones (*qnrB*), and sulfonamides (*sul*) (Supplemental Table S1). The incidence of the genes was previously reported in multiple studies ^16,55,56^ (Figure 4A). The *mcr* genes were observed independently in six isolates from BAPS2, BAPS3, and BAPS4 on small genomic fragments (Figure 4). We utilized long-read sequencing data to explore the genomic context of these genes. Among the 23 genes identified, six were found on plasmids and seven on both plasmid and chromosomal contexts. The ESBL gene *bla*_CTX-M_ was present on multiple plasmid backgrounds. The higher frequency of resistance genes in the BAPS1-6 groups is due to the presence of a range of plasmid-borne genes for aminoglycosides, β-lactams, tetracycline, florfenicol, fluoroquinolone, and sulfonamide resistance (Figure 4A). In recent clones, the BAPS1 group had a significantly higher content of resistance genes (p-value from proportion test < 0.05), due to the presence of plasmid-borne aminoglycoside modifying *ant3’’* and efflux pump *floR* genes in a higher number of isolates. The diversity of genomic contexts reveals multiple trajectories for the evolution of resistance, pointing to pervasive horizontal gene transfer of resistance genes and strong selective pressure from antimicrobial use in the poultry sector, resulting in the presence of these genes in recent clones ^57^. Despite the diversity of resistance genes, only a few appeared to be significantly linked to the resistance phenotype. For three classes of antimicrobials—aminoglycosides, β-lactams, and macrolides—we identified eight, three, and four resistance genes, respectively (Figure S2A). The most significant effects for aminoglycosides were observed for the aminoglycoside N-acetyltransferase (AAC) gene *aac3* and the aminoglycoside O-phosphotransferase (APH) gene *aph4*, with odds ratios significantly greater than one (Figure S2A, S2B). Both genes were plasmid-borne, accounting for 76% and 73% of the total resistance, respectively. Although the *ant3’’* gene also had a significant effect, it was smaller and found on both IncC plasmids and chromosomes (Figure S2A). For β-lactam resistance genes, the *bla*_CMY-2_ gene, present in 90% of isolates, significantly influenced the ampicillin resistance phenotype, accounting for 80% of the resistance phenotype, while its link with tazobactam was modest yet significant (odds ratio 2.3, resistance detection rate (RDR): 90%) (Figure S2A, S2B). The plasmid-borne *bla*_CTX-M_ gene, present in 20% of isolates, had a modest significant effect on tazobactam resistance. For macrolides, the *erm* and *mphA* genes, present in 2% and 3% of genomes, respectively, were significantly associated with azithromycin resistance (95% confidence interval of odds ratio was bigger than one), although they accounted for less than 30% of the resistance phenotype (Figure S2A, S2B, S2C).

**Figure 4:**
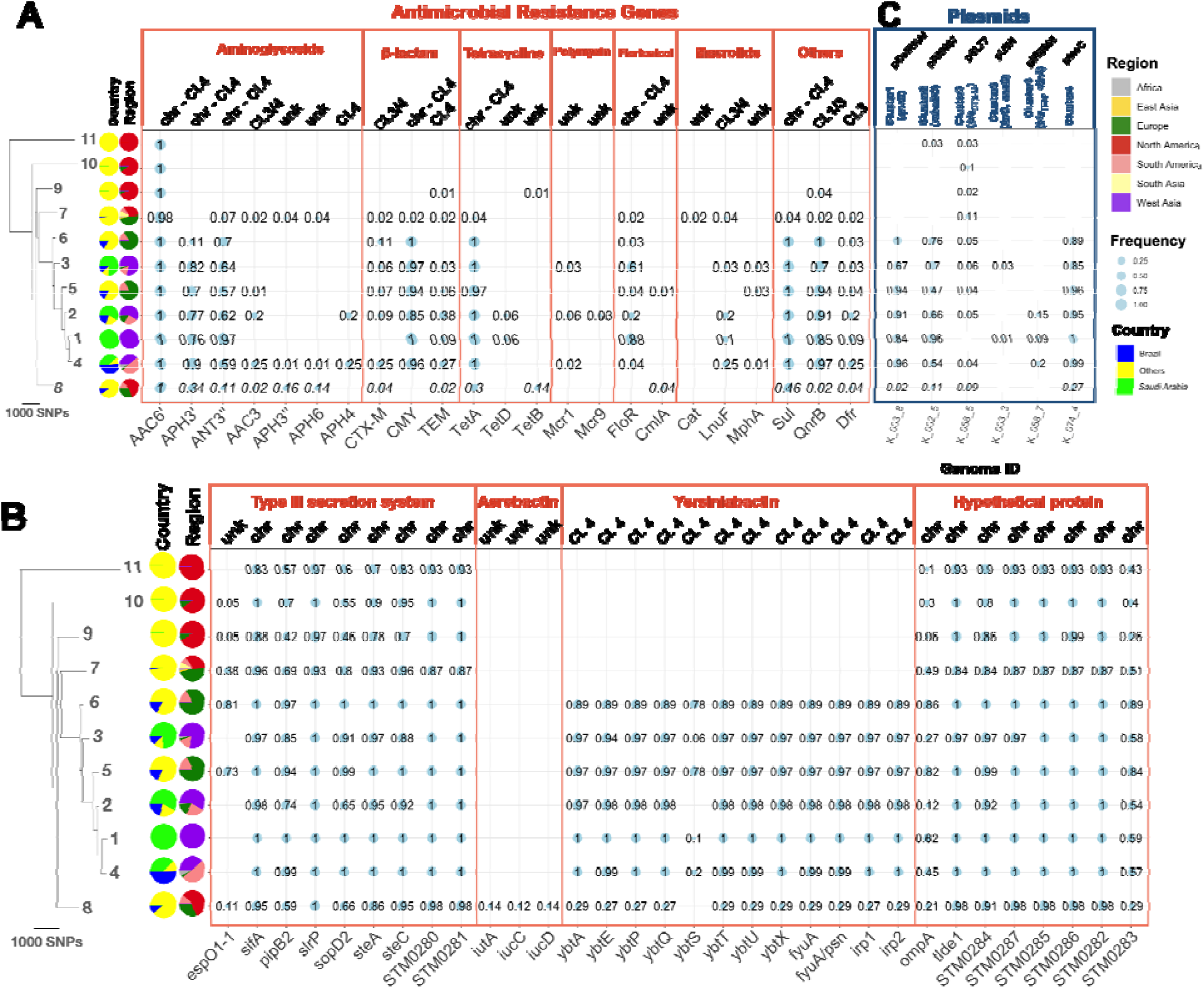
The distribution of **A)** antimicrobial resistance genes, **B)** virulence factor genes, and **C)** plasmids across the BAPS groups. Bubble sizes correspond to relative frequency, with numbers indicating relative frequencies within each BAPS group. The tree is the same as in Figure 1. For the detection of virulence factor genes and antimicrobial resistance genes, SRST2 with a threshold of 90% for identity and coverage was used. For plasmids in C), short reads for each strain were mapped against one of the plasmid backbones from each cluster presented in Figure S3A, using a cutoff of 90% coverage to determine the presence of the plasmid. For IncC plasmid, the shortest IncC plasmid backbone in the population was used as reference. The labels “chr” and “Cl.” denote “chromosomal” and “cluster,” respectively, indicating the genomic context of the gene on chromosomal or plasmid clusters as determined from examining the reconstructed plasmid/chromosomal contigs from long-read sequencing data. “Unk” denotes “unknown,” indicating cases where the genomic context could not be resolved due to either the absence of the genome in the collection or fragmented contigs.

### Variety of virulence mechanisms in *S.* Minnesota clones

*S.* Minnesota genomes were found to contain 213 distinct virulence genes, of which 88 genes were in the accessory genome, meaning they were present in less than 99% of strains. Among these 88 genes, 37 were distributed variably across the clones (Fisher’s test p-value < 0.05), suggesting the evolution of different pathogenicity mechanisms in the *S.* Minnesota clones. These genes included nine encoding components of the type III secretion system (T3SS) and eight less well-characterized genes, found on the chromosome (Figure 4B). All the variably present T3SS genes were effector proteins (Figure 4B), which, upon introduction across bacterial and host membranes, can disrupt host functions and are known to vary among pathogens ^58^. These genes may enhance invasion and adaptation in the primary host or the endurance of the strains in the environment, such as *sopD* in desiccation tolerance ^59^. Siderophore gene clusters of aerobactin and yersiniabactin were present exclusively in distinct clades. Aerobactin genes *iutA* and *iucCD* occurred in the BAPS8 group, while genes encoding the yersiniabactin operon (*fyuA, irp12, ybtAEPQSTUX*) were found in more recent BAPS groups (*ybtS* became detectable once the identity threshold was lowered to 0.6) (Figure 4B). The siderophore genes common to other *S.* Kentucky and APEC (Avian Pathogenic *Escherichia coli*) plasmids, including *iutA, iucABCD, sitABCD, etsABC, iss*, and *iroBCDEN* ^60^, were observed only in isolates from BAPS1-6 clones. While the virulence mechanisms in *S.* Minnesota are not fully understood, the acquisition of these iron chelator genes could facilitate *Salmonella* colonization and virulence in poultry, as previously observed in *S.* Heidelberg and *S.* Kentucky ^61,62^.

### Diversity of antimicrobial resistance genes carrying plasmids

We then characterized the plasmids in the clones by extracting all plasmid fragments with known plasmid replicons from the assemblies generated from long-read sequencing data. This analysis yielded 39 plasmid fragments, which were grouped into four clusters based on their sequence similarity (see Methods) (Figure S3A). Cluster 1 contained 3 Kb pRM plasmids, which include the plasmid mobilization genes *mbeBC*, necessary for plasmid transfer with the help of a conjugational plasmid, as well as a hypothetical protein (Figure S3A, S3B). These plasmids were more frequently found in the recent BAPS groups (47%-96%), significantly higher than in the ancestral BAPS groups (Figure 4C). Cluster 2 contained 3 Kb pColRNAI (Col(pHAD28)) plasmids, which carried the quinolone resistance efflux pump gene *qnrB1* (Figure 7B). This plasmid was present in most strains (67%-100%) of the emerging BAPS1 to 6 clones and was absent in the ancestral BAPS clones (Figure 4C). Cluster 3 included diverse plasmids, among which three carried resistance genes. One small pUWI plasmid (55 Kbp) harbored *linG*, an integron-associated gene cassette with an aminoglycoside adenyltransferase gene *aadA2*, which encodes a nucleotidyltransferase found in *Salmonella enterica*, conferring resistance to lincosamide antibiotics (Figure 7D). Another plasmid in this group, a small pIR plasmid (55 Kbp) reported in *Klebsiella pneumoniae* strains, carried β-lactam resistance gene *bla*_TEM_ and trimethoprim resistance gene *dfrA* (Figure S3D). Lastly, we identified a PSLT7 plasmid carrying the ESBL gene *bla*_CTX-M-8_, which possessed conjugation ability due to the presence of all transfer operon genes (Tra) (Figure S3D). These AMR-linked plasmids in cluster 3 were present at low frequencies in the recent BAPS groups 1-4 (1%-18%) (Figure 4C) and were sporadically acquired by lineages, which could lead to the emergence of novel clones with higher resistance levels from existing clones.

### The genomic configuration of IncC plasmids exhibits characteristics of mosaic plasmids, combining both resistance and virulence abilities

Cluster 4 includes 200Kb IncC plasmids that are large, low-copy, and frequently contain antimicrobial resistance (AMR) genes ^63^. These plasmids show mosaic structures with regions encoding both resistance and virulence genes, contributing to the higher resistance and virulence content in recent BAPS groups (e.g., BAPS 1-5) (Figures 8A and 5). Long-read sequencing data revealed that the largest IncC plasmids contain a virulence island composed of siderophore genes, including yersiniabactin *ybt, irp*, and *fyu* genes. Resistance genes are scattered across the backbone in several distinct locations, with most confined to a small region, as reported before (Figure 5A) ^63^. Six antimicrobial resistance genes were identified: *tet(A), aadA*, and *sul* were clustered together, while *bla*_CMY-2_, *aph(3)* and the ESBL *bla*_CTX-M-8_ were inserted separately (Figure 5A). The plasmid contains only some transfer (*tra*) locus genes, such as *traH* and *traF*, which might confer partial conjugative ability. Alignment with conjugative IncC plasmids from other Salmonella serovars (ST548 isolates Crie-F1017 and Crie-F1252), which are well-studied incompatibility group plasmids in *Salmonella* ^64^, confirmed the absence of several *tra* genes, suggesting conjugation may not be an inherent feature of the plasmids.

**Figure 5:**
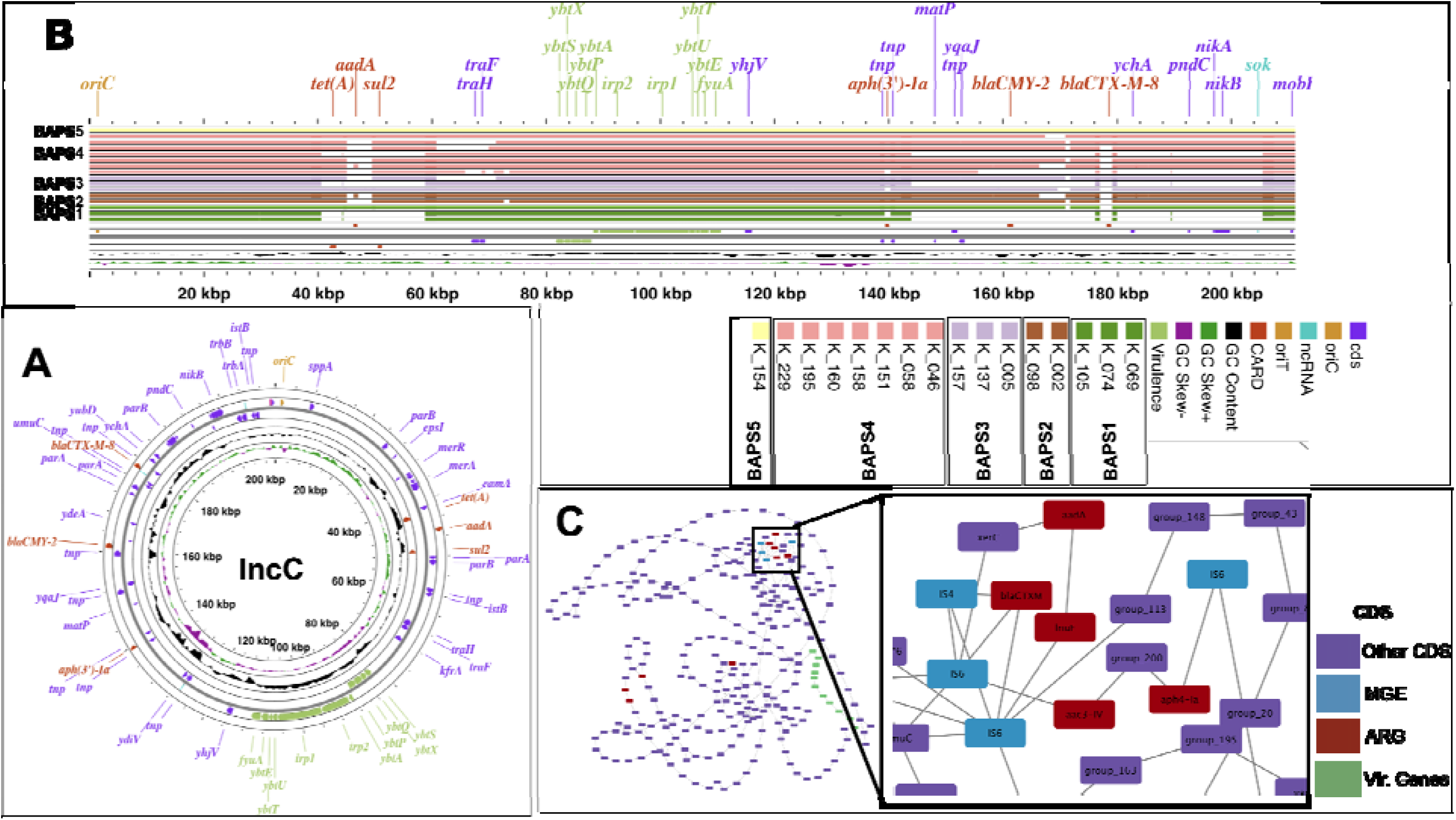
The structure and population genomics of IncC plasmids. **A)** Genomic map of the largest IncC plasmid (from isolate K_153) identified in the long-read sequencing data. **B)** Linearized IncC plasmid with alignment of all IncC plasmids against the plasmid in **A**. The bands for other plasmids are ordered from top to bottom and from left to right as indicated in the caption. Horizontal bars represent matches of 90% identity. Each color corresponds to the BAPS group from which the plasmid was isolated. **C)** Pangenome graph of IncC plasmids from 16 aligned IncC plasmids. Nodes and edges represent genes and their colocation within the same genomic context. Mobile genetic elements linked with resistance genes are colored blue.

### Genomic rearrangements and resistance gene dynamics in IncC Plasmids

IncC plasmids exhibited a variety of configurations across different strains. Among the 300 genes found in the plasmid fragments, 172 showed variable presence: 122 genes were present in 15% to less than 95% of genomes, and 50 genes were found in less than 15% of strains. Of these, 64 genes had functional annotations, including 25 mobile genetic elements and nine antimicrobial resistance genes. Regions containing resistance genes in IncC plasmids from strains in BAPS groups 1 to 5 displayed various rearrangements, as evidenced by multiple alignments of plasmid fragments (Figure 5B). In one plasmid (from isolate K_074), these changes resulted in the complete loss of entire resistance genes (Figure 5B). Further alignment at the gene-level also revealed rearrangements in the plasmid backbone due to mobilization of resistance genes, sometimes leading to inversion of entire virulence islands (Figure S4). The variation in gene order among plasmids within the same clone (BAPS groups 1 to 5) suggests that structural changes and recombination of plasmids occur concurrently with clonal expansion. The reconstructed synteny (pangenome) graph, where nodes represent genes and edges represent gene order, confirmed these findings. Resistance genes were predominantly situated in a highly interconnected part of the network, indicating numerous genomic rearrangements (Figure 5C). In contrast, virulence genes exhibited consistent gene order across plasmids (Figure 5C), highlighting that genomic diversity of IncC plasmids in expanding clones primarily originates from acquisition and loss of resistance genes.

## Discussion

We present the genomic analysis of a nationwide collection of *S.* Minnesota in the poultry chain, and revealed the population structure and diversity, especially the spread of antimicrobial resistance. Our results showed the emergence of new *S.* Minnesota clones with local and global distribution, formed over the past ten years. The epidemiology of S. Minnesota clones expanding both regionally and globally aligns with the import, export, and domestic production patterns of poultry meat, within Saudi Arabia. In 2020, Saudi Arabia produced 60% of its domestic poultry meat demand ^27^. Brazil remained the largest exporter of poultry meat to the country, while the UAE and other GCC countries were the main importers^27^. The presence of countries like South Africa and the UK, which are not direct importers or exporters of poultry meat from Saudi Arabia, in the transmission network for the identified clones suggests that the dissemination of *S.* Minnesota is facilitated by global poultry meat production and trade chains, beyond direct country-to-country or within-country exchanges.

Our study examines the largest collection of *S.* Minnesota in a consistent sampling framework and hence yielded accurate estimates of population parameters and dynamics of clones. Previous population genomics studies in the UK and Brazil on *S*. Minnesota traced the contamination to Brazil and showed a recent emergence of *S.* Minnesota clades ^10,65^. A study on a smaller collection of *S*. Minnesota in the UK traced the formation of *S.* Minnesota clades in the early 2000s, around the time of the introduction of the *S.* Enteritidis vaccine in Brazilian poultry in 2003 ^10^. Isolates from the UK from this study grouped with BAPS2 isolates, showing epidemiological links between the two collections. Our study also reports genetic relatedness between human and food production, although only one human sample was included. This potential link warrant a long-term surveillance study in both animal and human sectors could determine if the emergence of novel clades will pose significant risk to human health.

Our results underscore the key role of IncC plasmids in the recent evolution of *S*. Minnesota clones by providing a high-resolution population map of IncC plasmids. IncC plasmids pose a significant public health threat due to their broad host range and prevalence in animal, human, and environmental bacteria ^63,66^. The emergence of IncC plasmids in these reservoirs within enteric bacteria is associated with antimicrobial use in the agricultural sector [62]. Recent experimental works have shown that rearrangement of IncC plasmids in antimicrobial resistance encoding regions may occur in lab strains during the transfer process, which could enhance the fitness of the strains ^67–69^. The occurrence of different configurations of IncC plasmids within and between clones suggests plasmid recombination may serve as drivers for the adaptation in *S*. Minnesota in response to strong antimicrobial therapy in naturally occurring strains. Serovar switching and the rise of new clones and lineages have been ongoing in *Salmonella* populations over the past decades, attributed to multiple factors, including vaccines and antimicrobial treatment ^14^. The genomic dynamics of resistance genes of IncC plasmids, which co-occur with virulence genes on the same mosaic, may underpin the rise of novel clones from the *S.* Minnesota serovar with enhanced resistance and virulence.

Despite the breadth of the study, we highlight some limitations. Firstly, our collection mostly included multidrug-resistant strains. Although this allowed us to analyze multiple drug resistance profiles in depth, we could not assess the extent to which the emergence of new clones is driven by the selection for resistant strains. Moreover, the scarcity of samples from human sources prevented us from obtaining a full picture of the implication of the animal strains in human settings. Expanding the scope to include a broader collection, encompassing susceptible strains, will help determine whether the overall population structure of *S.* Minnesota remains consistent with our findings. Future studies may adopt the One Health appraoch, examining human, food, and animal strains within a unified consistent framework, to provide a comprehensive view of the diversification, virulence, and survivability of *S.* Minnesota strains. These studies will also enable the estimation of key parameters, including the rate at which the pathogen and antimicrobial resistance genes are introduced from food products into human settings. Such insights are essential for effectively treating, managing, and preventing illnesses in both humans and animals sectors ^70^.

## Disclaimer

The views expressed in this paper are those of the author(s) and not do not necessarily reflect those of the SFDA or its stakeholders. Guaranteeing the accuracy and the validity of the data is a sole responsibility of the research team.

## Authors contributions

Conceptualization was done by KOA, SMA, and DM. Methodology was developed by KOA, SMA, AAA, FMA, LEM, JH, DM, and GZ. Validation was carried out by KOA, SAA, AAA, and FMA. Formal analysis was conducted by JH and KOA. Research was conducted by SAA, AMA, SA, AAA, SIA, MSA, AA, FMA, ATA, NMA, AA, MA, TA, NA, JH, MM, EA, AAH, MA, and GZ. MB and DM provided resources. Data curation was performed by KOA, NMA, and LEM. KOA, DM and JH prepared the original draft. KOA, SMA, LEM and MAA contributed to writing, reviewing, and editing. JH handled visualization. SMA, FMA, KOA, and DM supervised the project, with project administration managed by SMA, KOA, and DM.

## Data Availability

https://www.ebi.ac.uk/ena/browser/view/PRJEB67737

