## Supplemental Figures for "Large-scale Genomic Survey of Non-typhoidal *Salmonella enterica* serovar Minnesota Strains in Chicken Products Reveals the Emergence of Multidrug Resistant Clones"

### Supplementary Figures

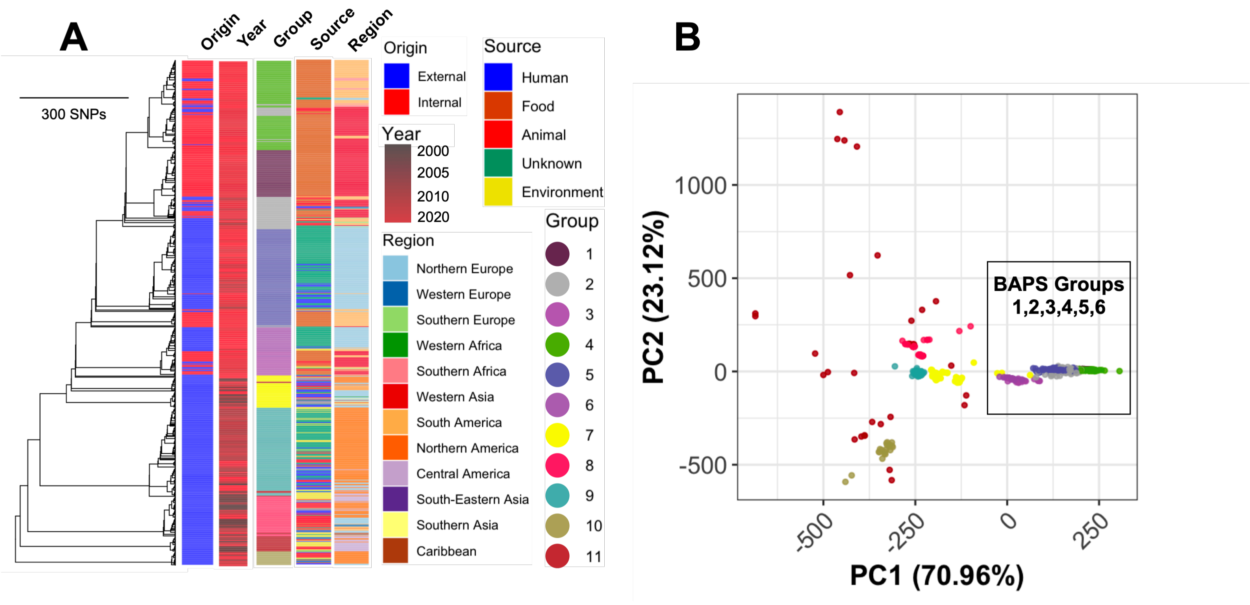


**Figure S1:** A) Whole genome sequencing tree based on 37,950 SNP sites for 734 strains across eleven BAPS groups. The tree is an approximately maximum likelihood phylogenetic tree rooted at the midpoint. The labels "internal" and "external" denote genomes from isolates studied here (originating from Saudi Arabia or Brazil) and genomes from the EnteroBase/PathogenDetection database, respectively. Branch lengths are scaled proportionally to sequence distances. B) Principal component analysis (PCA) plot representing the BAPS groups identified in the population based on pairwise genetic distance for the isolates in panel A. The colors of the BAPS groups correspond to the colors used in Panel A).

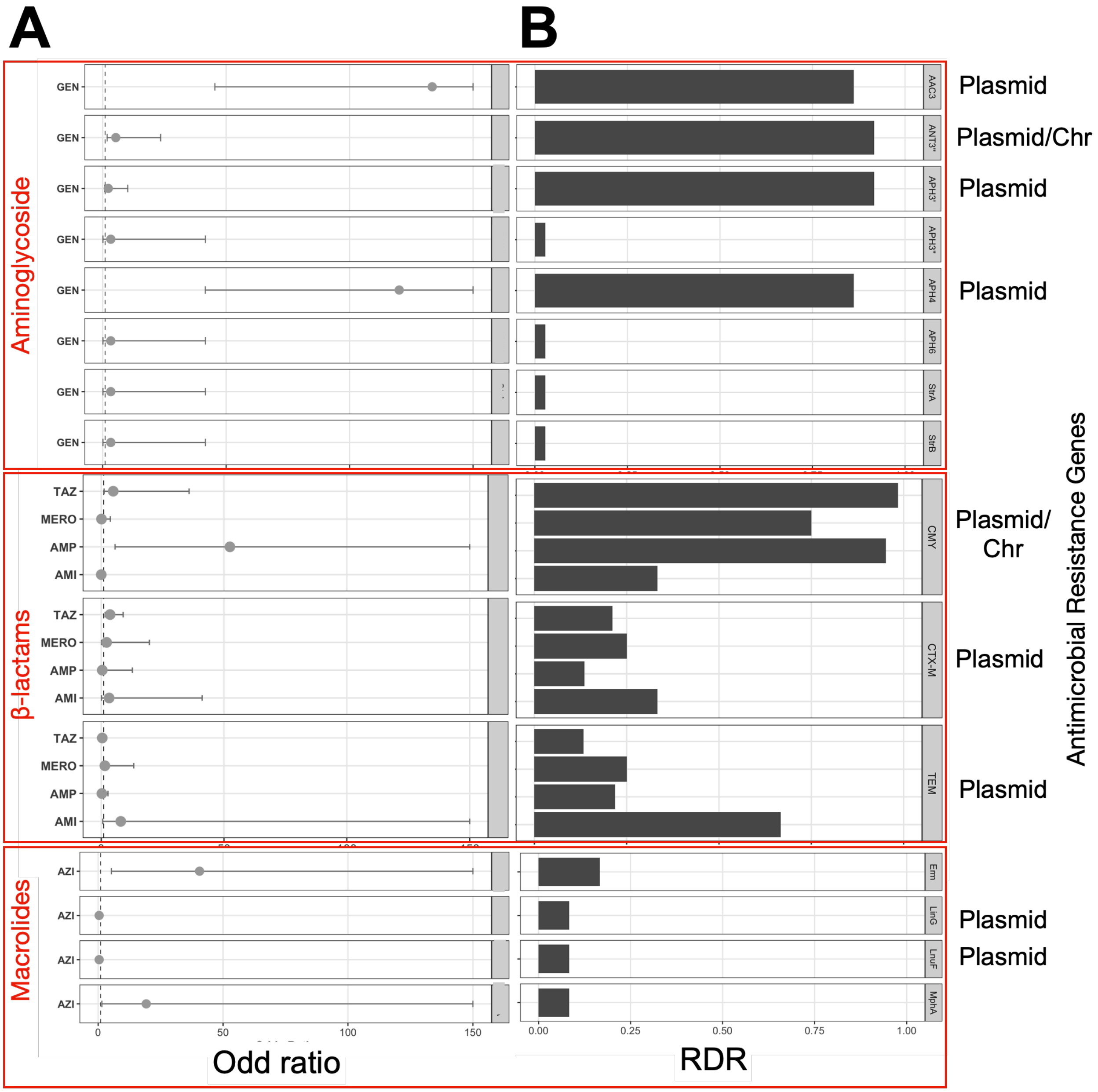


**Figure S2:** Integration of phenotypic and genotypic data. A) Odds ratio for the presence of resistance genes and their resistance phenotype, with error bars representing the 95% confidence interval. To improve clarity, the upper limit of the error bars was capped at 150. The labels "Plasmid" and "Chr" denote the plasmid and chromosomal genomic context of the gene from long-read sequencing data, respectively. B) Resistance detection rate (RDR), calculated as the number of strains with the resistant gene divided by the total number of resistant strains.

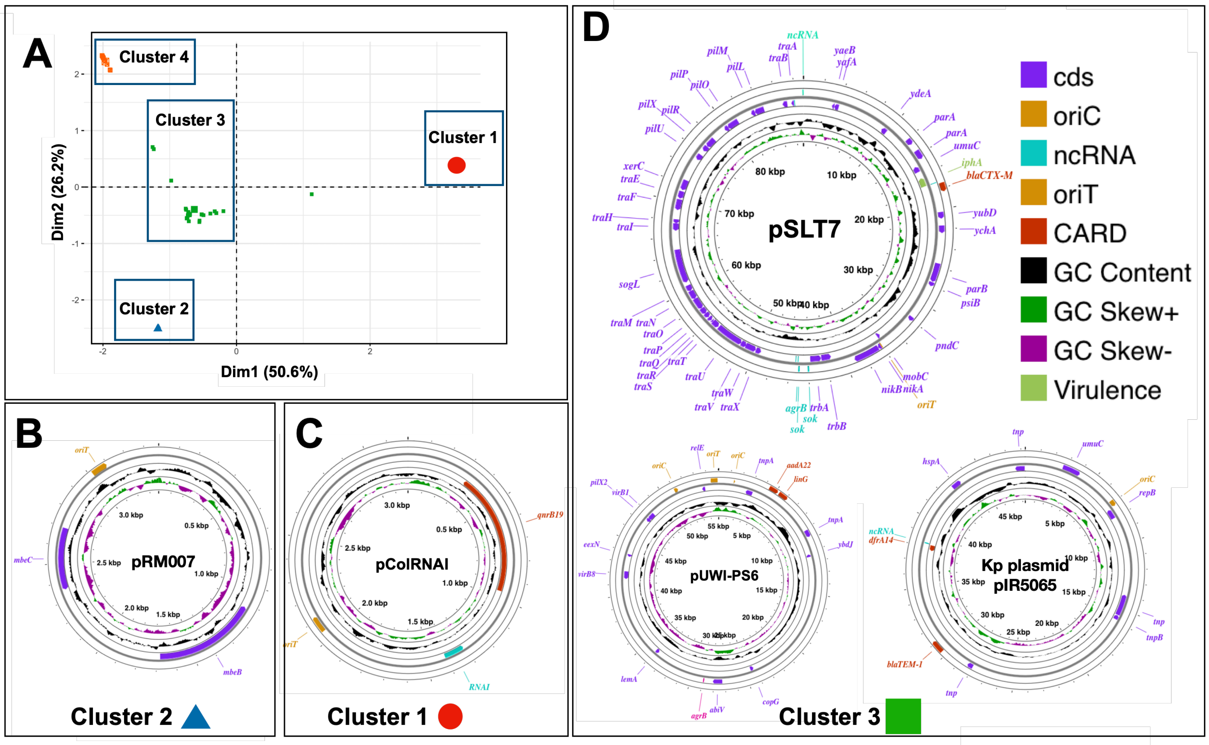


**Figure S3:** Diversity of plasmid fragments with known replicons in the population. A) Clusters of plasmids identified in the population through k-means clustering based on genetic distance between plasmid fragments (see Methods). B) Genetic map of representative plasmids containing known antimicrobial resistance genes and plasmid replicons. Plasmids are labeled according to the blast hit with the highest sequence identity.

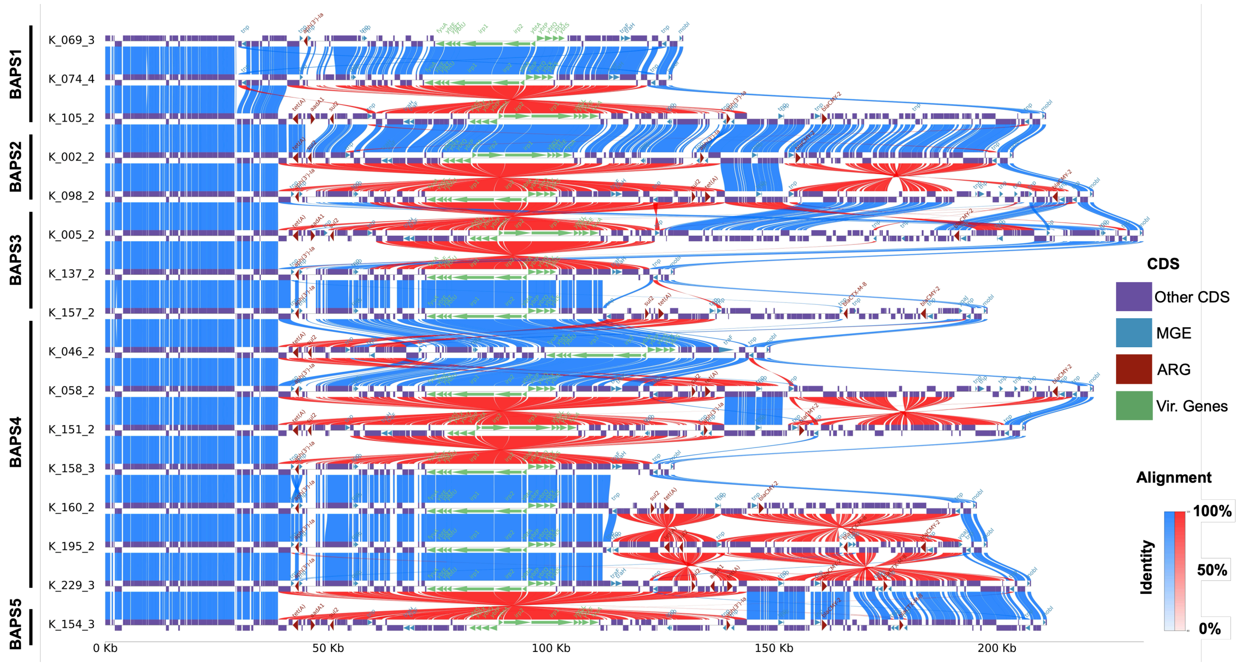


**Figure S4:** Genomic comparison (gene-level alignment) between IncC plasmids recovered from long-read sequencing data. Vertical lines indicate alignment, with color intensity reflecting sequence identity percentage. Red vertical lines denote inversion events. Horizontal bars represent coding sequence regions (CDS) for mobile genetic elements (MGE), antimicrobial resistance genes (ARG), and virulence factor genes (Vir. genes).
